# Elective Node Sparing in Head-and-Neck Cancer Radiotherapy Reduces Lymphocyte Damage, Lymphopenia, and Modulates Immune Signatures

**DOI:** 10.64898/2026.05.20.26352898

**Authors:** Justus Kaufmann, Ahmed Salah, Federico Marini, Sophia Drabke, Nina Gercek, Stephanie Breinich, Laura Oebel, Heinz Schmidberger, Sebastian Zahnreich

## Abstract

**Purpose:** Elective nodal (EN) irradiation (ENI) during radiotherapy for locally advanced head-and-neck squamous cell carcinoma (LA-HNSCC) influences hematotoxicity, anti-tumor immunity, and synergy with immunotherapy. We evaluated whether EN-sparing upfront boosts affect DNA damage, systemic immune signaling in peripheral blood lymphocytes (PBLs), and radiation-induced lymphopenia (RIL).

**Methods and Materials:** Twenty-eight patients with LA-HNSCC were randomized to either adjuvant or definitive chemoradiotherapy with standard ENI or EN-sparing upfront boost (adjuvant: 2×2 Gy; definitive: 5×2 Gy). Blood was collected pre-radiotherapy, 15 min, and 24 h after the first fraction, and before the sixth fraction. DNA damage in PBLs was assessed via γH2AX and 53BP1 foci and dicentric chromosome (DIC) assay. RNA sequencing was performed in two patients per group (definitive setting) at pre-CRT, before the sixth fraction, and at therapy end. Absolute lymphocyte counts (ALCs) were monitored weekly to assess RIL.

**Results:** DNA damage in PBLs correlated with planning target volume and whole-body dose, both of which were reduced by EN-sparing by 9.9-fold and 4.4-fold, respectively (p < 0.001 each). Correspondingly, EN-sparing significantly reduced radiation-induced foci and DIC levels in PBLs (≈3–4-fold, p < 0.001) and lowered the fraction of radiation-damaged PBLs per fraction (11% vs. 23% with ENI, p < 0.001). EN-sparing preserved baseline ALCs during week 1 of chemoradiotherapy and delayed RIL, whereas ENI caused an immediate ALC decline and RIL. Lymphocyte counts after week 1 negatively correlated with planning target volume, whole-body dose, and DNA damage in PBLs (p < 0.01). Transcriptomics showed metabolic and interferon signaling associated with EN-sparing, versus sterile inflammatory and damage-associated patterns with ENI.

**Conclusions:** EN-sparing by an upfront boost significantly reduced PBL damage and early RIL with distinct immune responses associated with lymphocyte viability and immune maturation. These findings support upfront EN-sparing strategies to mitigate RIL and improve radiotherapy–immunotherapy synergy in HNSCC.

## Introduction

Head and neck squamous cell carcinoma (HNSCC) is the sixth most common malignancy worldwide. In 2022, 946,456 new cases were diagnosed, and 482,001 patients died from this disease (1). Approximately 60% of patients present with locally advanced (LA) disease (stage III–IV), resistant to standard therapies and prone to recurrence and metastasis (2). Despite multimodal treatment, the 5-year overall survival for LA-HNSCC remains below 50% (2).

For more than two decades, the curative treatment paradigm for LA-HNSCC has remained largely unchanged. Resectable tumours are commonly managed by radical resection with neck dissection followed by adjuvant (chemo)radiotherapy (C)RT, while unresectable disease receives definitive CRT, occasionally preceded by induction chemotherapy (CT) (3). Many clinical trials have tested the addition of immune-checkpoint inhibitors (ICIs) targeting PD-1 or PD-L1 to improve outcomes. The majority of these trials failed to demonstrate a benefit; several large phase-III studies showed no improvement in disease control (4–7). The KEYNOTE-689 trial is the first in over 20 years to define a new standard of care: neoadjuvant ICI before surgery, followed by adjuvant CRT with continued ICI (8). Also, the NIVOPOST-OP trial (NCT03576417) showed improved disease-free survival by adding nivolumab to CRT in high-risk resected LA-HNSCC (9).

RT can enhance the efficacy of ICIs by inducing immunogenic tumor cell death, thereby promoting local and systemic antitumor immune responses (10). However, RT inevitably damages the radiosensitive hematologic system and can abolish the effect of ICIs in combined settings (11). Particularly, elective nodal (EN) irradiation (ENI) for LA-HNSCC has a significant negative impact on RT-ICI combinations (12, 13). Tumor-draining lymph nodes are key components of the cancer-immunity cycle, where tumor antigens are cross-presented and tumor-specific T cells undergo maturation (14). The upfront or complete exclusion of ENI can preserve RT-ICI synergy (12, 13, 15, 16). Current trials are investigating whether ENI for HNSCC without lymph node invasion should be omitted, de-escalated, or delayed (NCT03726775; ARO 2024-06) (17). The safety of reduced ENI has already been established (18), and ongoing clinical trials are investigating the effects of reduced or delayed ENI on the antitumoral immune response and RT-ICI efficacy (19).

Also, radiation-induced lymphopenia (RIL) is a common side effect of HNSCC RT, linked to poorer outcomes and may compromise RT-ICI efficacies (20). To date, the interplay between ENI and RIL remains largely unexplored (21). Besides simulations based on RT parameters to estimate dose to blood (22), biodosimetric markers of radiation-induced DNA damage in peripheral blood lymphocytes (PBLs) are used to assess blood exposure during RT (23). The most common assays detect γH2AX or 53BP1 DNA repair protein foci, as DNA double-strand break (DSB) surrogate markers, and chromosomal aberrations (23). More recently, RNA sequencing (RNA-seq) of blood has emerged as a high-resolution tool, enabling comprehensive profiling of transcriptional changes associated with radiation-induced tissue damage and immune modulation (24).

In this study, we examined how an upfront EN-sparing boost in definitive and adjuvant CRT for LA-HNSCC patients influences radiation damage, hematotoxicity, and systemic immune signaling, using radiation biomarkers in PBLs, RIL, and blood transcriptomics.

## Patients and Methods

### Patients and Therapy

This prospective, controlled, and randomized study included patients with pathological evidence of laryngeal, hypopharyngeal, p16-negative oropharyngeal, or oral cavity carcinoma, typical clinical symptoms, and a sufficient medical history. Patients who were unsuitable for surgical resection or declined surgery were considered for definitive CRT. Following surgery, patients with insufficient resection margins on pathological review or involved lymph nodes with extracapsular extension (ECE) were included for adjuvant CRT. RT planning and treatment were performed in accordance with local standards. For definitive therapy, a 5+5 approach was used (25), while EN volumes were defined according to international standards (26). In the adjuvant setting, volume and dose prescription also followed international standards (27). Boost volumes were defined based on pathological risk factors and could include lymph node levels with ECE, areas with close surgical margins, or both. In patients with definitive CRT, cisplatin was administered weekly at 40 mg/m² body surface area. In the adjuvant setting, cisplatin was administered with 20 mg/m² body surface area on five days in week one and week five of RT. The different treatment regimens and dose distributions for ENI versus EN-sparing are shown in Fig. 1. Both standard and time-adjusted equivalent dose in 2 Gy fractions (EQD2) values (28) for different treatment schedules are provided in the supplementary information (Appendix E1). Ethical approval was obtained from the Medical Association of Rhineland-Palatinate [No. 2024-17401 and 2024-17543]. All donors provided informed consent, and the research was conducted in accordance with the Declaration of Helsinki. RT was performed by 6 MeV volumetric modulated arc therapy with a Clinac DHX, Unique^TM,^ or Truebeam® (Varian Medical Systems Palo Alto, US) at the Department of Radiation Oncology and Radiation Therapy at the University Medical Center Mainz, Germany.

**Figure 1:**
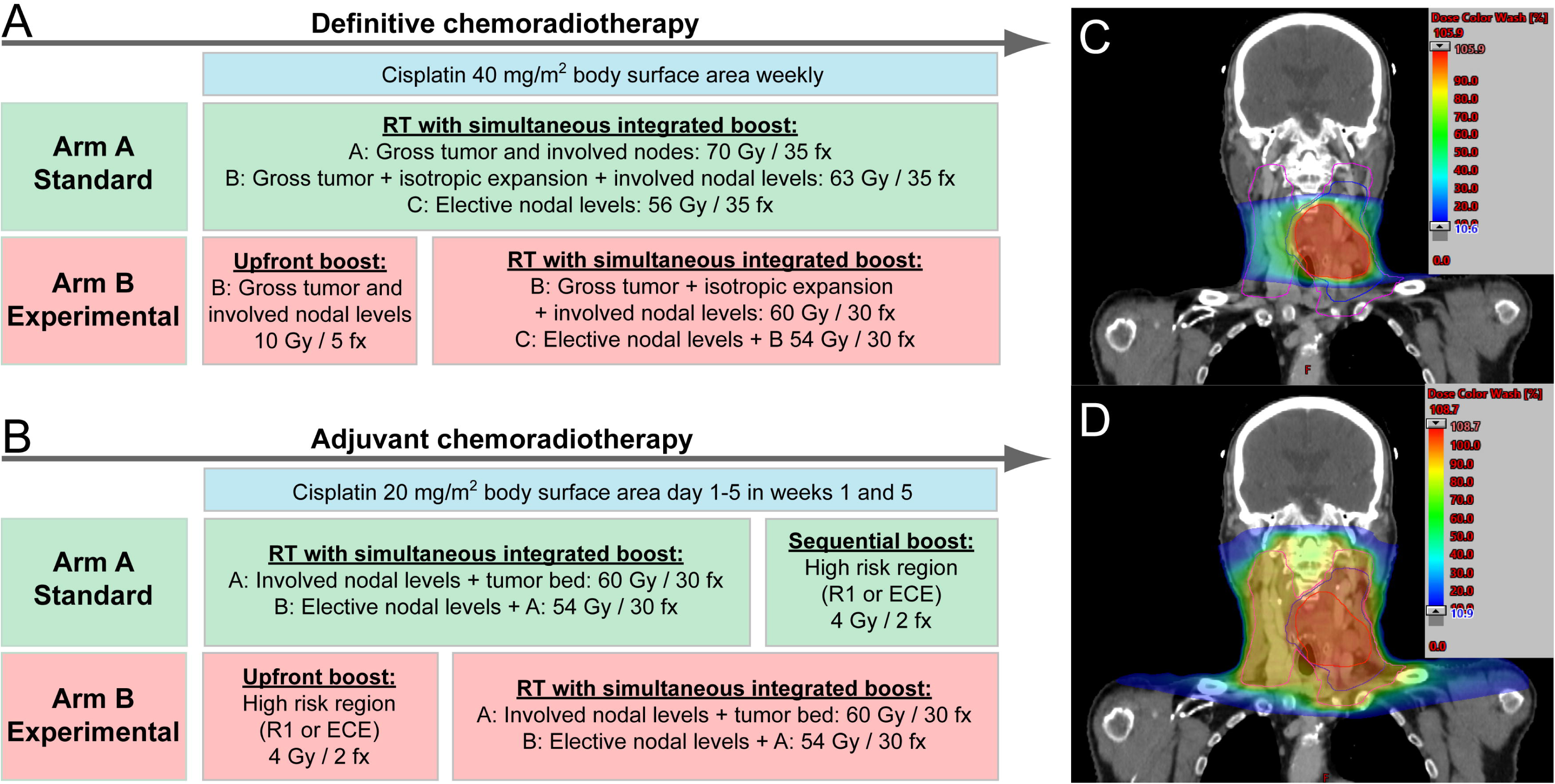
Treatment regimens for (A) definitive chemoradiotherapy (CRT) and (B) adjuvant CRT. At an alpha/beta of 10, doses were considered nearly isoeffective for definitive CRT patients using the equivalent dose in 2 Gy fractions (EQD2). Treatment planning images in dose color wash show a patient with a hypopharyngeal tumor in the experimental arm: (C) Elecitve nodal-sparing upfront boost and (D) after switching to elective nodal irradiation after 5 fractions. Planning target volumes are delineated as magenta: elective nodal levels, blue: high-risk nodal levels + primary tumor, red: macroscopic tumor. R1: positive surgical margins, ECE: extracapsular extension.

### Blood sampling and ex vivo irradiation

Venous blood collection, blood irradiation ex vivo, and isolation of PBLs were performed as described previously (23, 29). Blood was collected immediately before (pre-CRT), 15 min after, and 24 h after the first RT fraction. In definitive CRT, another sample was taken before the sixth fraction, and for transcriptomic analyses at the end of RT (end-CRT). Basal DNA damage was assessed in pre-CRT samples for all assays and subtracted from values after ex vivo or in vivo irradiation. A fraction of pre-CRT samples was irradiated ex vivo with 0.5 Gy and analyzed for γH2AX and 53BP1 foci 0.5 h post-irradiation.

### γH2AX and 53BP1 focus quantification

Ex vivo incubation, isolation, and fixation of PBLs, γH2AX and 53BP1 immunostaining, fluorescence microscopy, image capturing, and scoring of colocalizing γH2AX/53BP1 foci were performed as described previously (23, 30). In pre-CRT samples and all post-CRT samples, 500 cells were evaluated.

0.5 h after 0.5 Gy ex vivo irradiation of pre-CRT samples, 50 cells were analyzed. Radiation-induced foci (RIF) were assessed by subtracting foci in unirradiated pre-CRT samples from total post-irradiation values. Representative images for γH2AX/53BP1 foci in the supplementary information (Appendix E2).

### Dicentric Assay

The DIC assay was performed in blood samples from nine patients with definitive CRT obtained pre-CRT and before the sixth fraction as described previously (23). On average, 405 ± 138 (range: 203–631) metaphases were analyzed per sample. The basal yield of DIC in pre-CRT samples was subtracted from the corresponding post-RT samples.

### Whole-Body Dose

Each patient’s whole-body dose (WBD), as the average dose per voxel over the whole-body volume, was calculated as described previously (23). The distribution of the physical dose within the whole-body structure was calculated based on the data of the cumulative dose volume histogram (DVH) for the computed tomography-scanned body volume that was normalized to the whole-body volume. Treatment planning was conducted with “Eclipse” (Varian Medical Systems, Palo Alto, CA, USA).

### Lymphocyte Counts and Lymphopenia

Absolute lymphocyte counts (ALCs) were collected pre-CRT and subsequently every week during CRT. The National Cancer Institute’s Common Terminology Criteria for Adverse Events Version 5.0 (CTCAEv5.0) was used to classify treatment-related lymphopenia as grade 0/ normal: >1,000/mm^3^, grade 1: 800-999/mm^3^, grade 2: 500-799/mm^3^, grade 3: 200-499/mm^3^, and grade 4: <200/mm^3^ (31). Relative lymphocyte counts (RLCs) were calculated for each patient related to the pre-CRT baseline.

### Foci and DIC data and statistical analysis

Summarized average yields and data are provided as the mean and standard deviation (SD) unless stated otherwise. Data handling, plotting, and statistics were done using SigmaPlot (version 14, Systat Software, USA) and QtiPlot (version 5.12.8, IONDEV SRL, ROU). The relationship between two variables was analyzed using the Pearson test. For comparison of the means of two or more groups, the Student’s t-test or the one-way analysis of variance (ANOVA) was used, respectively. The chi-square test was used to evaluate associations between categorical variables. All levels of significance were set to α = 0.05.

### Blood transcriptomics

#### Samples and RNA Sequencing

Total RNA from blood samples was collected from two patients each with ENI or EN-sparing boost in the definitive setting pre-CRT, before the sixth fraction, and at end-CRT as described previously (29). One pre-CRT sample was unavailable for one patient with EN-sparing. Bulk RNA library preparation and transcriptome sequencing were conducted by Novogene (Novogene GmbH, Munich, Germany) according to the company’s protocols, and quality control was performed as reported previously (24).

#### Differential Expression and Gene Set Enrichment Analysis

Differential expression analysis was performed using the limma-voom framework on TMM-normalized counts after low-count filtering, retaining 19,129 genes (32). Genes on the X and Y chromosomes were removed before analysis to mitigate confounding by donor sex, which was collinear with treatment arm assignment in this cohort; pseudoautosomal region genes were retained given their autosomal expression behaviour. Three difference-in-differences (DID) contrasts were designated as primary analytical endpoints, comparing the change over time in the EN-sparing versus the ENI arm, using each group’s baseline as its own reference. DID isolated treatment-specific effects by accounting for baseline differences between groups, defined as: EN-sparing Δ (post-treatment − pre-treatment) − standard ENI Δ (post-treatment − pre-treatment). Moderated t-statistics were computed via empirical Bayes shrinkage. Gene set enrichment analysis (GSEA) on the moderated t-statistic ranking was used as the primary analytical output (33). GSEA was performed against the MSigDB Hallmark and Reactome collections. FDR < 0.05 was applied as the pathway-level significance threshold. Positive normalized enrichment scores (NES) in DID contrasts indicate greater enrichment in the EN-sparing arm.

## Results

### Patient and treatment characteristics

In total, 28 patients were included between April 2024 and December 2025 and were block randomized into two groups of CRT ±ENI for either adjuvant or definitive treatment. 15 patients received definitive CRT (+ENI: n = 9, −ENI: n = 6), and 13 patients adjuvant CRT (+ENI: n = 6, −ENI: n = 7). RT was discontinued prematurely in one case due to patient preference. All other patients received full-dose RT. Concurrent CRT was administered in 22 patients. One patient received weekly docetaxel due to an age of over 80. All other patients received cisplatin, and 13 patients received a cumulative cisplatin dose of at least 200 mg/m² body surface area. Patient characteristics are summarized in Table 1. Sex was not used to stratify patients, resulting in an imbalance between study arms: 71.4% of women (5/7) and 38.1% of men (8/21) were treated with an EN-sparing boost.

### Basal levels and ex vivo radiosensitivity

To assess basal DNA damage markers and radiation response in patient PBLs, we measured background γH2AX/53BP1 foci and DIC in unirradiated pre-CRT samples, and RIF 30 minutes after 0.5 Gy ex vivo irradiation. Individual levels of DNA damage in PBLs and raw data are provided in the supplementary information (Appendices E2 and E3). Basal γH2AX/53BP1 foci per PBL varied markedly among patients (0.37 ± 0.18, range: 0.04–0.88, CV = 48%), highlighting the need to account for variability after irradiation. After 0.5 Gy X-rays, mean RIF counts per PBL were similar after basal correction (4.40 ± 0.33, range: 3.67–4.93, CV = 7.4%), indicating comparable DSB induction across patients. Basal DIC were detected only in two of 9 analyzed patients scheduled for definitive CRT and were subtracted from post-CRT data.

### Elective nodal-sparing reduces DNA damage in peripheral blood lymphocytes

Fifteen minutes after the first RT fraction, all patients exhibited RIF in PBLs, strongly correlating with planning target volume (PTV) size and WBD (Fig. 2A). Individual RIF levels during RT and corresponding raw data are in the supplementary information (Appendices E3 and E4). EN-sparing reduced PTV and WBD by 9.9-fold and 4.4-fold compared to standard ENI (1,304 ± 351 cm³ versus 132 ± 60 cm³, p < 0.001; 93 ± 21 mGy versus 21 ± 9.0 mGy, p < 0.001, each). Accordingly, mean RIF frequency per PBL was about 4-fold significantly lower with EN-sparing (0.87 ± 0.39 versus 0.23 ± 0.13 RIF per cell; Fig. 2A), indicating concordance between reductions in WBD and RIF per PBL.

**Figure 2:**
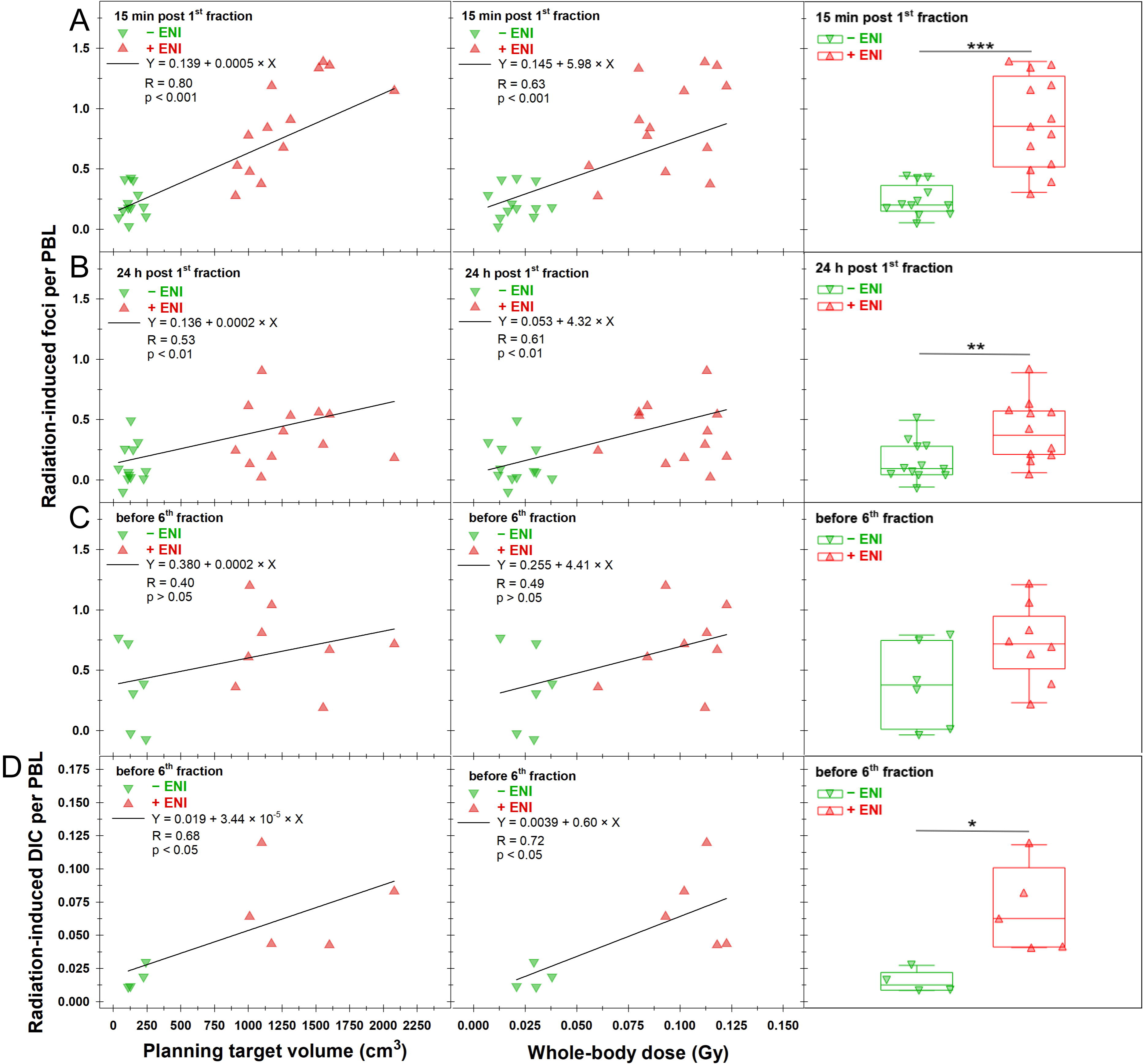
Radiation-induced (A-C) γH2AX/53BP1 foci and (D) dicentric chromosomes (DIC) in patients’ peripheral blood lymphocytes (PBLs) after radiotherapy. Data are shown relative to planning target volume (left panel), whole-body dose (middle panel), and stratified by elective nodal irradiation status (± ENI, right panel) (A) 15 min after the first fraction, (B) 24 h after the first fraction, and (C, D) before the sixth fraction (definitive only). The lines in the left and middle panels represent linear fits to the data, along with the corresponding function, Pearson correlation coefficient (R), and the p-value of the fit. Statistical comparison between chemoradiotherapy ± ENI (right panel) was conducted using Student’s t-test (* p < 0.05; ** p < 0.01; *** p < 0.001).

At 24 h after the first fraction, 83% of patients (19/23) showed a reduction of initial post-RT RIF per PBL (Fig. 2B). Patients with ENI, initially with higher RIF levels post-RT, exhibited an average 2.4-fold decline to 0.36 ± 0.21 RIF per PBL (p < 0.01). In contrast, only 67% of EN-sparing patients showed an on average 1.7-fold reduction to 0.14 ± 0.17 RIF per PBL (p < 0.05). A moderate correlation between residual RIF frequency and both PTV size and WBD persisted at 24 h, with mean residual RIF per PBL remaining significantly 2.8-fold higher in the standard ENI group than in the EN-sparing group (0.39 ± 0.25 versus 0.14 ± 0.17 RIF per PBL; Fig. 2B).

In 14 patients undergoing definitive CRT, RIF levels were measured before the sixth fraction, prior to adaptation to ENI, to assess RIF accumulation during CRT between treatment arms (Fig. 2C). Higher RIF levels before the sixth fraction versus 24 h after the first fraction were seen in 63% (5/8) of ENI patients and 67% (4/6) of EN-sparing patients. In contrast, compared to initial levels 15 min after the first fraction, only 25% (2/8) with ENI and 50% (3/6) with EN-sparing showed higher residual RIF. At this time, RIF per PBL still showed moderate but non-significant correlations with the PTV and WBD. The mean level of residual RIF was about 2-fold higher with ENI versus EN-sparing but not significant (0.71 ± 0.23 versus 0.36 ± 0.36 RIF per PBL, Fig. 2C). However, RIF accumulation in PBLs during RT is strongly influenced by DSB repair, fractionation intervals, and treatment interruptions, making direct comparison between study arms at this time point imprecise. To enable robust comparison, the DIC assay was used since DICs accumulate during RT, providing a more reliable marker of cumulative RT-induced DNA damage (23). DIC frequencies in PBLs correlated strongly with PTV size and WBD (Fig. 2D). Mean DIC frequency per PBL after EN-sparing was significantly 3.7-fold lower than with ENI (0.019 ± 0.009 versus 0.072 ± 0.032), consistent with the ∼4-fold difference in initial RIF 15 min after the first fraction.

To determine the proportion of RT-damaged PBLs and the distribution of DNA damage within this population, we performed dispersion analyses of RIF post-RT. The fraction of radiation-damaged PBLs with ≥1 RIF correlated strongly with PTV size and WBD 15 min after the first fraction (Fig. 3A). The average proportion of radiation-damaged PBLs was significantly 2.1-fold lower in patients with EN-sparing compared with ENI. Patients with ENI showed an overall significantly 13% higher proportion of radiation-damaged PBLs compared to EN-sparing (10.9 ± 6.5% versus 23.3 ± 9.8%, Fig. 3B), associated with significantly higher proportions of PBLs with multiple RIF (2–15 per cell), reflecting more severe and potentially lethal RT-induced damage. 24 h after the first fraction and before the sixth fraction, only weak correlations between the fraction of radiation-damaged PBLs and the PTV or WBD were observed, and overall differences between the treatment groups were no longer significant (see supplementary information, Appendix E5).

**Figure 3:**
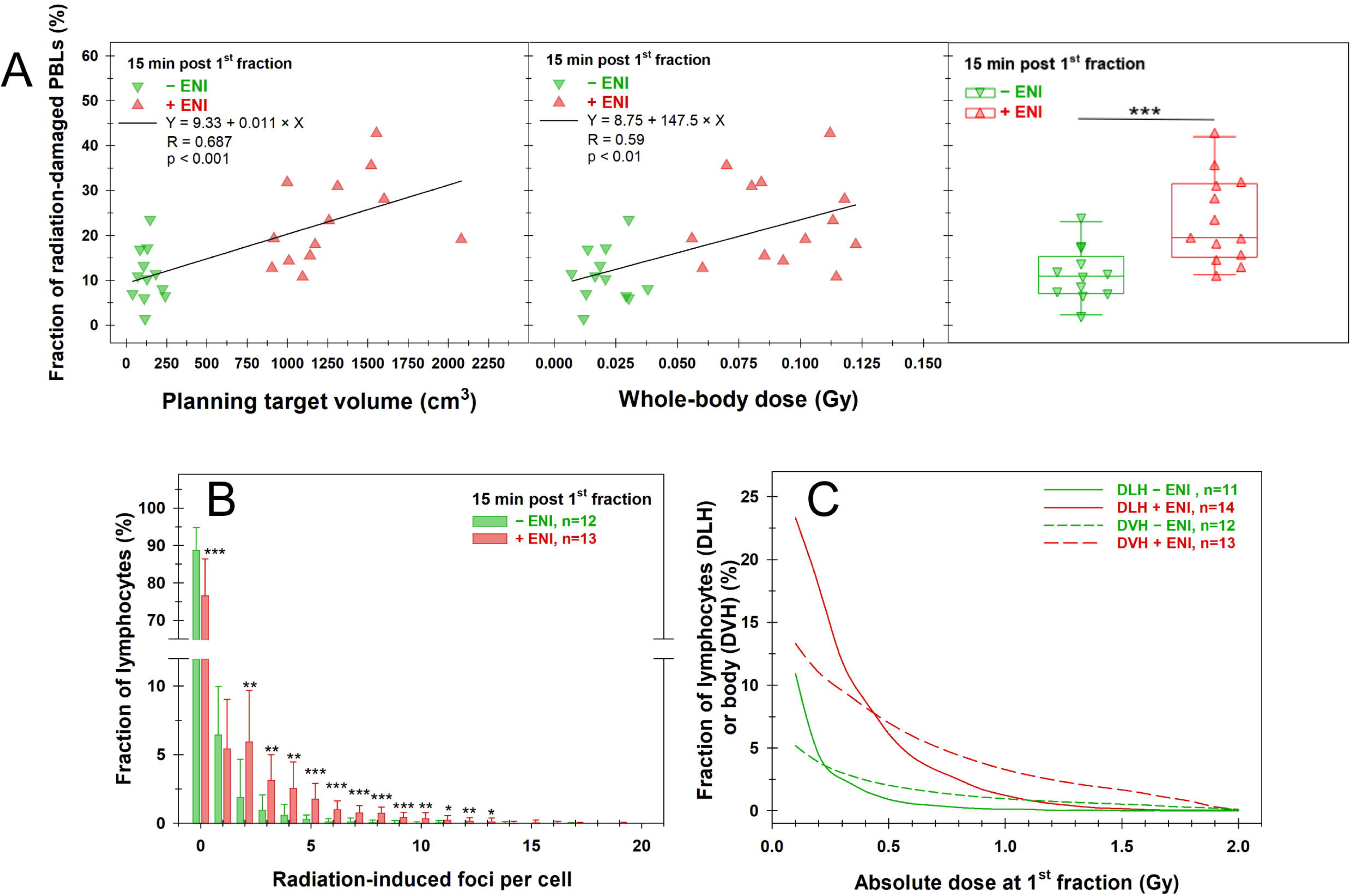
Analysis of the proportion of radiation-damaged peripheral blood lymphocytes (PBLs) and the distribution of biological damage indicators and physical dose distribution in patients. (A) Fraction of PBLs with ≥1 radiation-induced γH2AX/53BP1 focus after radiotherapy relative to planning target volume (left panel), whole-body dose (middle panel), and stratified by elective nodal irradiation status (± ENI, right panel) 15 min after the first fraction. The lines in the left and middle panels represent linear fits to the data, along with the corresponding function, Pearson correlation coefficient (R), and the p-value of the fit. (B) Distribution of radiation-induced γH2AX/53BP1 foci in patients’ PBLs 15 min after the first fraction of radiotherapy ± ENI. (C) Cumulative dose–lymphocyte histograms (DLH) and dose–volume histograms (DVH) derived from the proportion of exposed lymphocytes 15 min after the first fraction or the total body volume. Data are presented as the mean and standard deviation. Statistical comparison between radiotherapy ± ENI was conducted using Student’s t-test (* p < 0.05; ** p < 0.01; *** p < 0.001).

For comparison with the dose distribution in the body volume predicted by the cumulative DVHs of the treatment planning software, cumulative dose-to-lymphocyte histograms (DLHs) were generated, based on RIF dispersions 15 min after the first fraction. Conversions from average RIF per PBL into dose were based on our previously published reference data, using a dose-response of 10 RIF per PBL per Gy (23). While the integral DLHs and partial body volume based on DVHs did not align, they reflected the differences in dose distribution between CRT with or without ENI. However, compared with the DVH data for partial-body exposure, DLH analysis generally overestimated low-dose exposure and underestimated higher-dose exposure, particularly in the ENI group.

### Elective nodal-sparing preserves peripheral lymphocytes and delays lymphopenia

After the first week of CRT, EN-sparing did not significantly change ALCs relative to the pre-CRT baseline in either the adjuvant or definitive setting, whereas ENI caused an immediate drop (Fig. 4A). A comparable decline in ALCs was observed in the EN-sparing group in week 2 after adjustment for delayed ENI.

**Figure 4:**
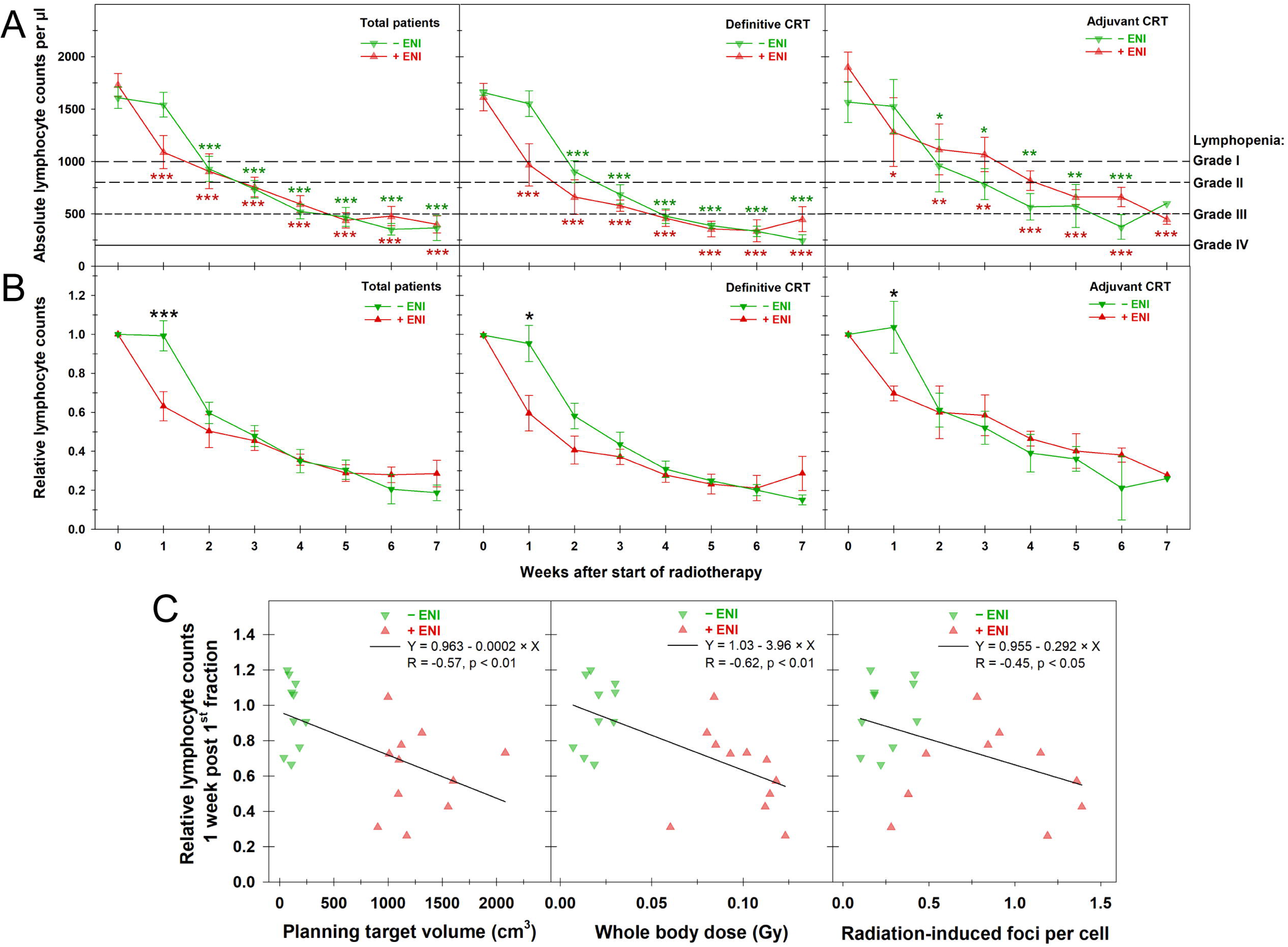
Lymphocyte counts during chemoradiotherapy (CRT). (A) Absolute or (B) relative lymphocyte counts (RLCs) for total patients (left panel) and for patients receiving definitive (middle panel) or adjuvant (right panel) CRT stratified by elective nodal irradiation status (± ENI). Grades of lymphopenia per CTCAE v5 are indicated. Data are presented as the mean and the standard error of the mean. The patient counts for each data point are in the supplementary information (Appendix E6). Statistical comparison between (A) pre-CRT and post-CRT within each treatment group or (B) between CRT ± ENI was conducted by One-Way ANOVA or Student’s t-test, respectively (* p < 0.05; ** p < 0.01; *** p < 0.001). (C) Correlations of relative lymphocyte counts per patient after one week of CRT and the planning target volume (left panel), whole body dose (middle panel), and radiation-induced foci per cell 15 min after the first fraction (right panel). The lines represent linear fits to the data, along with the corresponding function, Pearson correlation coefficient (R), and the p-value of the fit.

The onset and severity of RIL are summarized in Table 2. All patients developed RIL during therapy; however, onset timing differed significantly between treatment arms (χ² = 9.78, df = 4, p < 0.05). In the ENI cohort, RIL occurred in week 1, whereas in the EN-sparing arm, it first appeared in week 2. Notably, the week 2 incidence in the EN-sparing group closely matched the cumulative incidence by week 2 in the ENI group (58% versus 61%, 7/12 versus 7/13). Beyond week 2, the temporal evolution of RIL was comparable between groups. Regarding RIL grade at nadir, no statistically significant difference was observed between treatment arms. Although a higher proportion of patients in the standard ENI group developed grade IV RIL (21% versus 8%, 3/14 versus 1/13), this difference was not statistically significant.

Due to pronounced inter-patient variability in ALCs, RLCs were used for the comparison between treatment arms (Fig. 4B). For all patients, and when stratified by definitive or adjuvant CRT, RLCs remained unchanged during the first week of CRT for EN-sparing and were significantly higher compared to the ENI group. Individual patient data for ALC and RLC are in the supplementary information (Appendix E6). RLCs after week 1 exhibited significant negative correlations with the PTV, WBD, and the number of RIF per PBL 15 min after the first fraction (Fig. 4C).

### Blood transcriptome profiling reveals progressive immune divergence between treatment arms

To examine hematologic systemic therapy responses at the molecular level, bulk RNA-seq data from blood samples were analyzed. Pre-CRT, the treatment arms showed minimal autosomal transcriptomic differences following sex gene removal (67 genes at nominal *p* < 0.01 versus ∼191 genes expected under null distribution), supporting baseline comparability for longitudinal comparison. DID analysis revealed distinct transcriptomic profiles during CRT between treatment arms. Before the sixth fraction, the EN-sparing group showed significantly greater enrichment of lymphocyte metabolic and proliferative gene programs, including oxidative phosphorylation (NES = +2.70, FDR < 0.001), E2F targets (NES = +2.13, FDR < 0.001), MYC targets (NES = +1.96, FDR < 0.001), DNA repair (NES = +1.96, FDR < 0.001), and glycolysis (NES = +1.88, FDR < 0.001). Conversely, in the ENI arm, TNF-α signaling via NFκB was significantly elevated (NES = −1.52, FDR = 0.012), indicating early onset of damage-associated inflammatory signaling.

At end-CRT, DID analysis across CRT showed greater type I interferon (IFN-α; NES = +2.39, FDR < 0.001) and type II interferon (IFN-γ; NES = +1.65, FDR = 0.001) enrichment in the EN-sparing arm, alongside continued elevation of oxidative phosphorylation (NES = +3.12, FDR < 0.001) and MYC targets (NES = +2.47, FDR < 0.001). In the ENI group, dominant mitotic spindle gene enrichment (NES = −2.61, FDR < 0.001), TGF-β signaling (NES = −1.72, FDR = 0.013), and IL6/JAK/STAT3 signaling (NES = −1.36, FDR = 0.022) were observed.

Within-arm trajectory analysis confirmed these divergent fates: the EN-sparing arm showed strong upregulation of IFN-α (NES = +2.86, FDR < 0.001) and IFN-γ (NES = +2.39, FDR < 0.001) from pre-CRT to end-CRT, while the ENI arm showed dominant inflammatory (NES = +2.19, FDR < 0.001) and TNFα activation (NES = +1.98, FDR < 0.001) without commensurate type I IFN upregulation (IFN-α, p > 0.05) (Fig. 5A and B). Cross-sectional comparison at end-CRT corroborated these findings, with the EN-sparing group showing higher IFN-α response (NES = +2.31, FDR < 0.001) and oxidative phosphorylation (NES = +2.41, FDR < 0.001), and the ENI group showing higher inflammatory response (NES = −1.68, FDR = 0.002), TNF-α signaling (NES = −1.75, FDR < 0.001), and mitotic spindle enrichment (NES = −2.59, FDR < 0.001) (Table 3).

**Figure 5:**
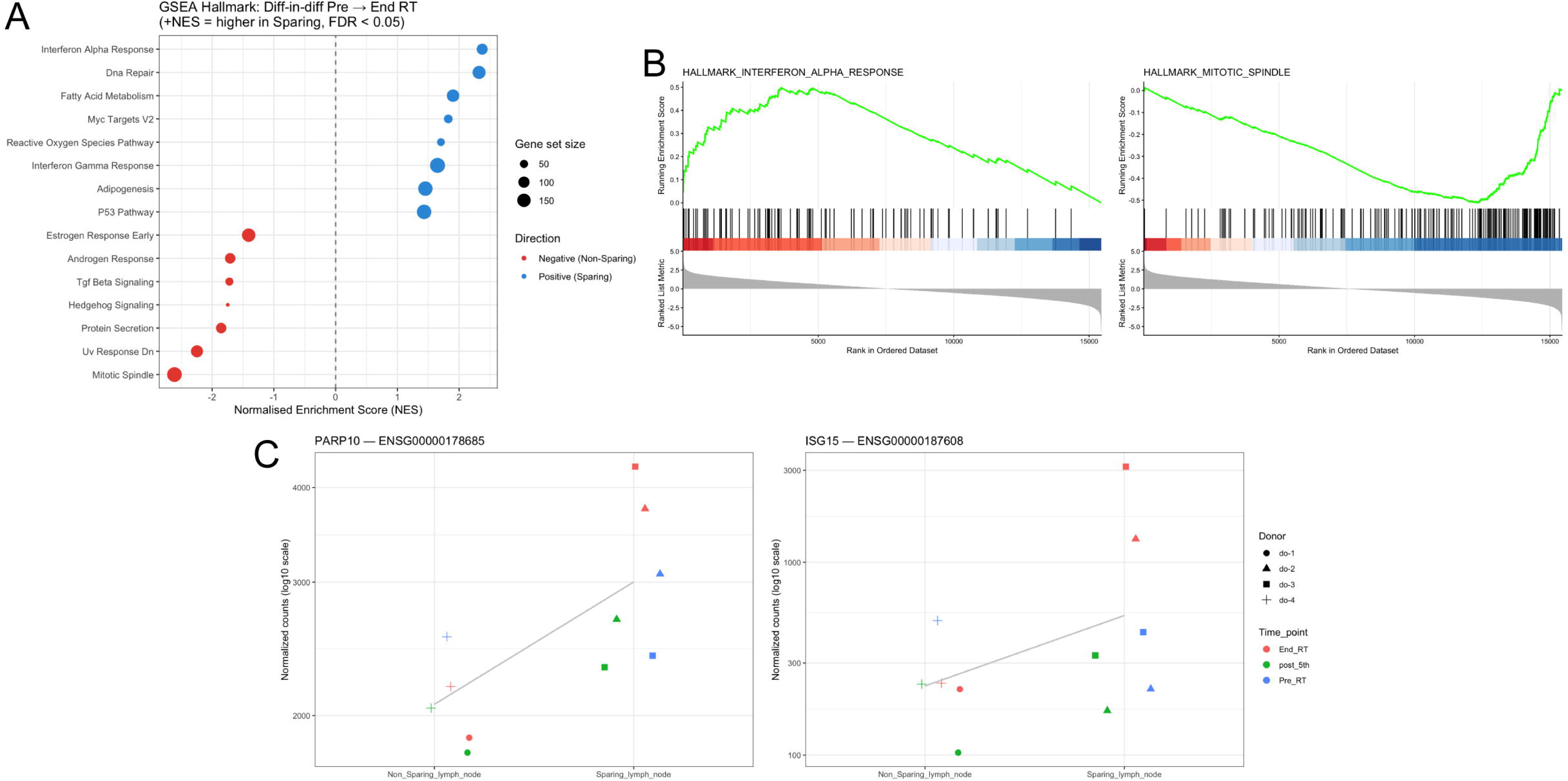
Radiotherapy effects on the transcriptomic response in whole blood. (A) Gene set enrichment analysis (GSEA) comparing the pre-chemoradiotherapy (CRT) to end-CRT trajectory in the elective nodal (EN)-sparing versus the elective nodal irradiation (ENI) arm. Only pathways reaching a false discovery rate (FDR) < 0.05 are shown. Positive normalized enrichment scores (NES) indicate greater enrichment in the EN-sparing arm; negative NES indicate greater enrichment in the ENI arm. (B) GSEA barcode plots for Interferon-α Response (NES = +2.39, FDR < 0.001), enriched in the EN-sparing arm, and Mitotic Spindle (NES = −2.61, FDR < 0.001), enriched in the ENI arm. The ranked list metric (bottom panel) reflects the moderated t-statistic from the difference-in-differences contrast. (C) Normalised expression counts of ISG15 (left panel) and PARP10 (right panel) across treatment arms and timepoints. Each point represents one sample; donor identity is encoded by shape and timepoint by colour.

At the individual gene level, several genes upregulated specifically in the EN-sparing group at end-CRT confirmed the pathway-level findings (Fig. 5C). ISG15, a canonical IFN-stimulated gene, was among the differentially expressed genes, providing direct gene-level support for the IFN-α pathway enrichment observed in this group. PARP10, a MYC-interacting protein involved in replication fork maintenance during active DNA synthesis, was also elevated, consistent with the MYC, E2F, and DNA repair program signatures and reflecting replication-coupled activity in proliferating lymphocytes.

## Discussion

This study examined how an upfront EN-sparing boost in definitive and adjuvant CRT for LA-HNSCC patients influences hematotoxicity and systemic immunogenic signaling, using radiation biomarkers and transcriptomic analysis in PBLs, as well as assessing effects on RIL. To our knowledge, this is the first study to explore upfront EN-sparing boost strategies in LA-HNSCC across both treatment settings with these methods.

Boost strategies, delivering extra doses to the primary tumor as sequential or simultaneous integrated boosts, are commonly used in HNSCC and affect radiobiological outcomes and toxicity (34). Sequential boost concepts at the end of treatment, with lower biologically effective doses to EN regions, have mainly been explored in dose de-escalation to reduce toxicity in HPV-positive HNSCC (35). Alternatively, an upfront EN-sparing boost may preserve lymphoid structures, reduce RIL, and thereby enhance T-cell priming, antitumor immune memory with sustained immune response (12).

For DNA damage marker induction in PBLs, we found good to very good correlations with PTV and the WBD for RIF up to 24 h after the first fraction and for DIC before the sixth fraction, consistent with our prior findings in other tumor types after partial-body irradiation (23). This led to a significant reduction in the mean DNA damage frequencies per PBL due to EN-sparing with smaller PTVs and WBDs. Accordingly, the fraction of radiation-damaged PBLs with RIF immediately after the first fraction correlated strongly with both PTV and WBD. We observed a significantly lower average proportion of 13% radiation-damaged PBLs per RT-fraction with EN-sparing. The higher proportion of radiation-damaged PBLs in patients with ENI was associated with increased counts of RIF per PBL. Overall, given the high radiosensitivity of PBLs, a higher rate of cell death via apoptosis can be expected (30, 36). When the mean RIF per PBL was converted to dose using our reference data of 10 RIF per cell per Gy (23), average values 15 min after the first fraction corresponded to 0.087 and 0.023 Gy per PBL for ENI and EN-sparing, respectively. Considering only RIF in radiation-damaged PBLs, doses were higher, averaging 0.53 and 0.21 Gy per affected PBL for ENI and EN-sparing, respectively. Our previous studies on radiosensitivity of isolated PBLs showed 14% apoptotic cells at 0.25 Gy and 22% at 0.5 Gy (30).

Based on the 1.6-fold higher induction of apoptotic cells at the mean dose in radiation-damaged PBLs between treatment arms, we estimate theoretical apoptosis rates of ≈6.4% for EN-sparring and ≈27.2% for ENI after five RT fractions. Predicted viable lymphocyte fractions are therefore ≈94% and ≈73% of baseline. These align very well with empirical findings: mean RLC after week 1 of CRT was 99% ± 18% in the EN-sparring group versus 63% ± 23% in the ENI group. This close match suggests RIF in PBLs strongly correlates with apoptosis rate and RIL. The significant negative correlation between the rates of RIFs in PBLs and the RLC also confirmed this, indicating that RIF in PBLs after RT could serve as a rapid, predictive biomarker for RIL. Previously, only Lee et al. (37) showed a moderate correlation between the fraction of PBLs with chromosomal aberrations and the nadir of normalized white blood cell and lymphocyte counts in 22 lung cancer patients at the end of C-ion RT. However, RIL was mild due to normal tissue sparing with particle RT. Because of the small patient sample, we found no correlation between DIC yields in PBLs or the fraction of aberrant PBLs and RLCs after the first week of CRT.

In this study, all patients developed RIL during CRT. About 40% of ENI patients showed RIL after week 1 of CRT, whereas RIL with EN-sparing did not emerge until week 2, notably despite only a 2-fold upfront EN-sparing boost in the adjuvant setting. This highlights the detrimental impact of large-volume ENI on the peripheral blood system, beyond direct damage to nodal regions. By week 3, nearly all patients in both arms showed RIL. Under ENI, slightly more patients reached grade 4, and most total patients reached at least grade 3. Even if comparable RIL occurred later, the early benefit of EN-sparing may still allow initial antitumor T-cell priming in EN, antitumor T-cell expansion, and immunological memory formation, as supported by our transcriptomic data. However, patient numbers in both study arms were small, limiting statistical power and generalizability. While trends suggest a less pronounced severe immunotoxic effect with EN-sparing after the onset of RIL, larger prospective cohorts are needed to confirm these findings and assess clinical relevance, particularly for long-term immune function and tumor control.

In general, the severity of RIL depends on RT parameters like target volume, number of fractions, and dose per fraction (38). Also, pre-RT ALC and irradiation of large blood vessels and lymphoid organs contribute to overall and persisting RIL (39–41). We also demonstrated negative correlations between RLCs and PTV, with even stronger correlations for WBD. The latter finding suggests that incorporating additional patient-specific parameters improves the predictive value of the immunotoxic impact of RT on RIL. Although computational approaches for modeling blood dose during RT exist (22), biomarker evaluation in PBLs offers direct in vivo measurements that incorporate patient-specific (radio)biological characteristics.

So far, few studies have examined the impact of ENI on RIL in HNSCC patients. Only recently, a dose-response relationship for EN de-escalation trials in definitive HNSCC CRT and RIL was established by Mickel et al. (21). In their study, ENI treatment involved 40 Gy in 20 fractions to involved and adjacent nodal levels only, no ENI, or standard-of-care with ENI. The extent of ENI exposure significantly impacted grade 4 RIL at nadir, and up to 9 months post-RT. As our study adopted ENI after the fifth or second upfront boost fraction in the definitive or adjuvant setting, and RIL was only monitored during CRT so far, direct comparison is not feasible. However, with ENI from the outset, more patients reached grade 4 RIL in our study, and long-term follow-up will provide additional data.

RIL has also been considered a potential cause of failure of RT-ICI combinations during HNSCC RT/CRT (42–45). The majority of studies indicate that pre-treatment ALCs in HNSCC do not affect the outcome of subsequent ICI therapy, whereas persistent RIL represents a negative prognostic factor. Particularly, since curative CRT for HNSCC can perturb crosstalk between lymphoid and myeloid compartments, driving long-term shifts in immune cellular composition that may impair antitumor immunity and reduce ICI efficacy (46).

To identify differences in cellular signaling pathways and potential biomarkers across treatment groups, we conducted transcriptomic analysis of patient blood. We observed distinct immunological responses after the first week and different trajectories throughout CRT using a DID framework. The early metabolic activation signature in the EN-sparing group up to the sixth fraction is consistent with the observed preservation of ALC. In contrast, ENI caused early damage-associated inflammatory signaling, driven mainly by innate myeloid and stromal cells responding to damage-associated molecular patterns from irradiated tissue (47), rather than lymphocyte-mediated adaptive immunity, consistent with an immediate ALC drop. Signaling pathways linked to CRT course, evident by treatment end, include type I/II IFN signaling in the EN-sparing group, while the ENI group showed higher general inflammatory activity, TNF-α signaling, and mitotic spindle enrichment. The observed EN-sparing sequence, where metabolic divergence precedes IFN divergence, aligns with lymphocyte biology: early antigen-driven activation and metabolic reprogramming precede lymphocyte maturation and effector T-cell expansion associated with IFN secretion (48). The mitotic spindle enrichment in the ENI group indicates emergency myelopoiesis associated with a higher damage burden and loss of blood cells (49). Similarly, IL-3/GM-CSF signaling drives the survival and expansion of myeloid progenitors, and chronically elevated myeloid cytokine signaling is associated with myeloid-derived suppressor cell differentiation (50). Crucially, this inflammatory, myeloid-dominant program in the ENI group occurred without corresponding type I IFN activation. Together, the key findings were the distinction between IFN-driven adaptive immunity in the EN-sparing group, associated with antiviral and antitumor immunity, and damage-driven sterile inflammation in the ENI arm.

## Conclusion

Our study shows that upfront EN-sparing boost strategies in LA-HNSCC significantly reduce RT-induced DNA damage in PBLs, delay RIL, and promote transcriptomic profiles associated with systemic long-term antitumor immunity. Prospective validation in larger cohorts is needed to confirm whether reduced early peripheral lymphocyte depletion with EN sparing improves the distribution of antitumor-primed T cells, antitumor memory, and ultimately ICI efficacy and patient outcomes.

## Data Availability Statement for this Work

Research data are stored in an institutional repository and will be shared on request to the corresponding author. Processed RNA-seq data are available at https://github.com/AhmedSAHassan/HNSCC_Transcriptomic_Dataset

## Acknowledgements

We thank all patients participating in this study, Giusy Carlino for technical lab assistance, and Antonina Thomaidis for her support as a study nurse.

