## supplementary information for "Elective Node Sparing in Head-and-Neck Cancer Radiotherapy Reduces Lymphocyte Damage, Lymphopenia, and Modulates Immune Signatures"

### Appendix E1

Time-dependent equivalent dose in 2 Gy fractions (EQD2) was calculated considering tumor repopulation starting with a delay of 28 days and 0.9 Gy for every day thereafter. EQD2 was calculated for both normal tissue (α/β= 3) and tumor (α/β = 10). For normal tissue, the dose could be considered almost identical, with less than 0.5 Gy difference in total dose. Time-dependent EQD2 suggests at least similar efficacy when considering tumor repopulation.

| Dose calculation | Elective nodal irradiation arm | Elective nodal-sparing arm | Intermediate risk: elective nodal irradiation arm | Intermediate risk: Elective nodal-sparing arm |
|---|---|---|---|---|
| EQD2 (α/β = 3) | 51.52 | 51.84 | 60.48 | 60 |
| EQD2 (α/β = 10) | 54.13 | 53.1 | 61.95 | 60 |
| Time dependent EQD2 (α/β = 10) | 39.88 | 45.47 | 53.05 | 60 |

### Appendix E2

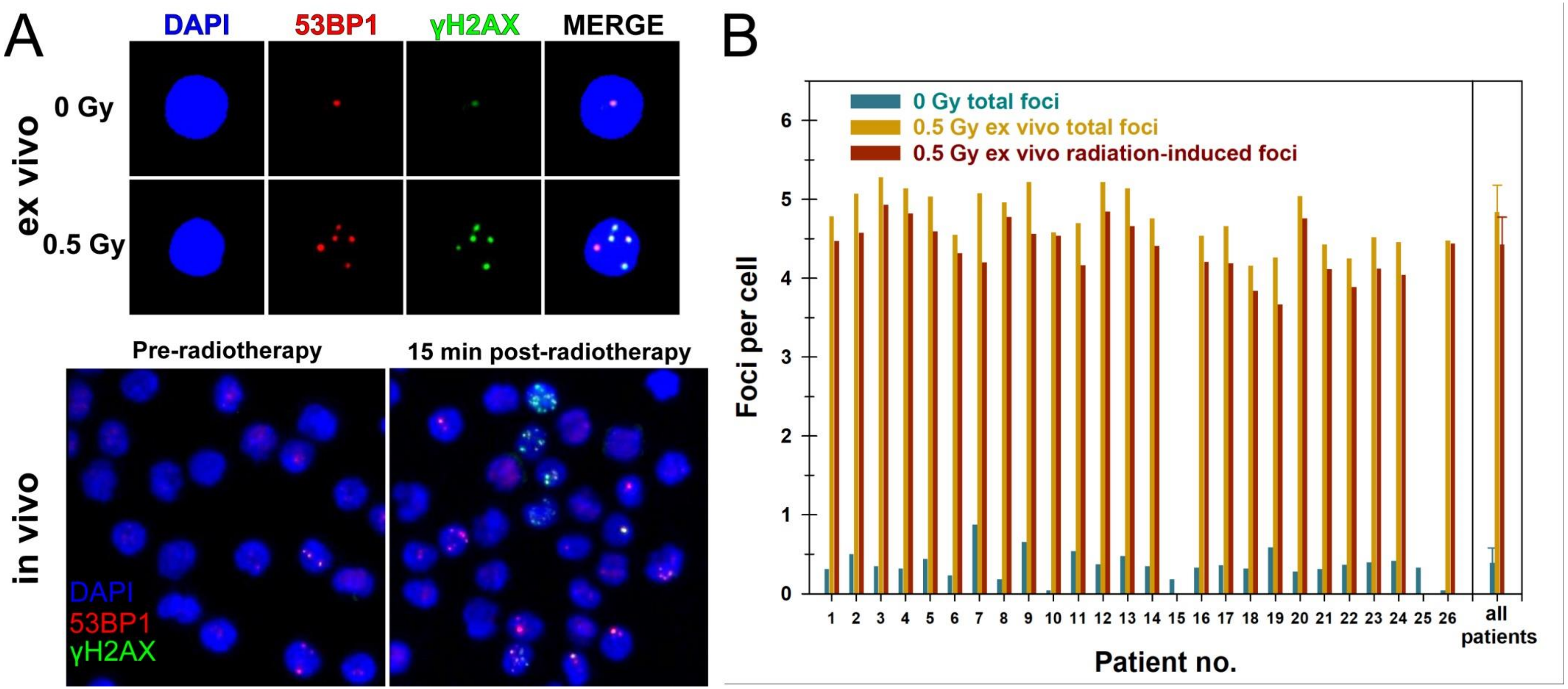


Quantification of colocalizing γH2AX/53BP1 foci in peripheral blood lymphocytes (PBLs). (A) Representative fluorescence microscopy image of basal (0 Gy) and ex vivo radiation-induced (0.5 Gy, 30 min) foci in PBLs as quantified (upper panel). Overview fluorescence image of foci in PBLs before and 15 minutes after the first radiotherapy fraction (lower panel). (B) Individual basal and ex vivo radiation-induced (0.5 Gy, 30 min) foci in PBLs collected before radiotherapy from each patient. Shown are total foci counts for 0 Gy and 0.5 Gy ex vivo and only radiation-induced foci (0.5 Gy ex vivo – 0 Gy).

### Appendix E3

Individual and summarized data for the planning target volume (PTV), whole-body dose (WBD), total and radiation-induced (RI) γH2AX/53BP1 foci per peripheral blood lymphocyte (PBL), proportions of total and radiation-damaged PBLs harboring at least one RI γH2AX/53BP1 focus per cell, and total and RI dicentric chromosomes.

| | | | | | | | | | | | | | | | | | | | | | | | |
|---|---|---|---|---|---|---|---|---|---|---|---|---|---|---|---|---|---|---|---|---|---|---|---|
| | End-point | | | | Foci per cell | | | | | | | | | Fraction cells with yH2AX/53BP1 foci (%) | | | | | | | Dicentrics per cell | | |
| | Iradiation | | | | sham (0 Gy) | ex vivo 0.5 Gy | | in vivo | | | | | | sham (0 Gy) | in vivo | | | | | | sham (0 Gy) | in vivo | |
| | Time point post-exposure | | | | $t_1$ (30 min) | | | $t_2$ (15 min post 1. fx) | | $t_3$ (24 h post 1. fx) | | $t_4$ (before 6. fx) | | $t_1$ (30 min) | $t_2$ (15 min post 1. fx) | | $t_3$ (24 h post 1. fx) | | $t_4$ (before 6. fx) | | $t_1$ (30 min) | $t_4$ (before 6. fx) | |
| | Setting | Pat. no. | PTV (cm3) | WBD (Gy) | Total | Total | RI | Total | RI | Total | RI | Total | RI | Total | Total | RI | Total | RI | Total | RI | Total | Total | RI |
| + ENI | definitive | 1 | 1009 | 0.093 | 0.31 | 4.78 | 4.47 | 0.79 | 0.48 | 0.45 | 0.14 | 1.52 | 1.21 | 23.1 | 37.51 | 14.41 | 30.42 | 7.32 | 59.20 | 36.10 | 0.0000 | 0.0654 | 0.0654 |
| | | 3 | 1601 | 0.118 | 0.35 | 5.28 | 4.93 | 1.71 | 1.36 | 0.90 | 0.55 | 1.03 | 0.68 | 35.90 | 64.07 | 28.17 | 40.51 | 4.62 | 50.10 | 14.20 | 0.0000 | 0.0441 | 0.0441 |
| | | 4 | 1171 | 0.123 | 0.32 | 5.14 | 4.82 | 1.51 | 1.19 | 0.52 | 0.20 | 1.37 | 1.05 | 42.32 | 60.36 | 18.04 | 34.00 | -8.32 | 60.60 | 18.28 | 0.0000 | 0.0449 | 0.0449 |
| | | 5 | 2080 | 0.102 | 0.44 | 5.04 | 4.60 | 1.59 | 1.15 | 0.63 | 0.19 | 1.17 | 0.73 | 27.94 | 47.15 | 19.21 | 32.00 | 4.06 | 57.46 | 29.51 | 0.0526 | 0.0844 | 0.0844 |
| | | 16 | 1553 | 0.112 | 0.33 | 4.54 | 4.21 | 1.72 | 1.39 | 0.63 | 0.30 | 0.53 | 0.20 | 23.60 | 66.40 | 42.80 | 40.00 | 16.40 | 31.60 | 8.00 | N/A | | |
| | | 20 | 1099 | 0.113 | 0.28 | 5.04 | 4.76 | N/A | | 1.19 | 0.91 | 1.10 | 0.82 | 16.97 | N/A | | 52.19 | 35.23 | 48.43 | 31.46 | 0.0000 | 0.121 | 0.121 |
| | | 23 | 904 | 0.060 | 0.40 | 4.52 | 4.12 | 0.68 | 0.28 | 0.65 | 0.25 | 0.77 | 0.37 | 21.40 | 34.22 | 12.82 | 32.18 | 10.78 | 37.08 | 15.68 | N/A | | |
| | | 26 | 999 | 0.084 | 0.04 | 4.48 | 4.44 | 0.82 | 0.78 | 0.66 | 0.62 | 0.66 | 0.62 | 1.80 | 33.65 | 31.85 | 35.20 | 33.40 | 36.35 | 34.55 | N/A | | |
| | adjuvant | 7 | 1121 | 0.085 | 0.88 | 5.08 | 4.20 | 1.72 | 0.84 | N/A | | N/A | | 48.70 | 64.29 | 15.58 | N/A | | N/A | | N/A | | |
| | | 12 | 1572 | 0.080 | 0.37 | 5.22 | 4.85 | 1.71 | 1.34 | 0.94 | 0.57 | | | 27.15 | 62.77 | 35.62 | 49.00 | 21.85 | | | | | |
| | | 14 | 916 | 0.056 | 0.35 | 4.76 | 4.41 | 0.88 | 0.53 | N/A | | | | 24.15 | 43.53 | 19.38 | N/A | | | | | | |
| | | 19 | 1728 | 0.113 | 0.59 | 4.26 | 3.67 | 1.27 | 0.68 | 1.00 | 0.41 | | | 22.20 | 45.60 | 23.40 | 49.00 | 26.80 | | | | | |
| | | 24 | 1191 | 0.0802 | 0.42 | 4.46 | 4.04 | 0.80 | 0.38 | 0.45 | 0.03 | | | 26.19 | 37.05 | 10.86 | 28.63 | 2.44 | | | | | |
| | | 25 | 1312 | 0.080 | 0.33 | N/A | | 1.24 | 0.91 | 0.87 | 0.54 | | | 21.40 | 52.40 | 31.00 | 42.32 | 20.92 | | | | | |
| | n | | 14 | 14 | 14 | 13 | 13 | 13 | 13 | 12 | 12 | 8 | 8 | 14 | 13 | 13 | 12 | 12 | 8 | 8 | 5 | 5 | 5 |
| | Mean | | 1304 | 0.093 | 0.39 | 4.82 | 4.42 | 1.26 | 0.87 | 0.74 | 0.39 | 1.02 | 0.71 | 25.92 | 49.92 | 23.32 | 38.79 | 14.62 | 47.60 | 23.47 | 0.0105 | 0.0720 | 0.0720 |
| | SD | | 351 | 0.021 | 0.18 | 0.34 | 0.37 | 0.42 | 0.39 | 0.23 | 0.25 | 0.34 | 0.33 | 11.18 | 12.44 | 9.79 | 8.00 | 13.40 | 11.34 | 10.66 | 0.0235 | 0.0321 | 0.0321 |
| | Min | | 904 | 0.056 | 0.04 | 4.26 | 3.67 | 0.68 | 0.28 | 0.45 | 0.03 | 0.53 | 0.20 | 1.80 | 33.65 | 10.86 | 28.63 | -8.32 | 31.60 | 8.00 | 0.0000 | 0.0441 | 0.0441 |
| | Max | | 2080 | 0.123 | 0.88 | 5.28 | 4.93 | 1.72 | 1.39 | 1.19 | 0.91 | 1.52 | 1.21 | 48.70 | 66.40 | 42.80 | 52.19 | 35.23 | 60.60 | 36.10 | 0.0526 | 0.121 | 0.1210 |
| | CV (%) | | 26.9 | 22.81 | 47.41 | 7.01 | 8.35 | 33.10 | 44.61 | 31.67 | 64.14 | 33.85 | 46.51 | 43.16 | 24.91 | 42.00 | 20.63 | 91.63 | 23.83 | 45.41 | 223.61 | 44.54 | 44.54 |
| - ENI | definitive | 2 | 110 | 0.0303 | 0.50 | 5.07 | 4.57 | 0.68 | 0.18 | 0.57 | 0.07 | 1.23 | 0.73 | 28.50 | 34.66 | 6.16 | 14.67 | -13.83 | 52.37 | 23.87 | 0 | 0.0125 | 0.0125 |
| | | 9 | 127 | 0.0208 | 0.66 | 5.22 | 4.56 | 0.84 | 0.18 | 0.69 | 0.03 | 0.65 | -0.01 | 37.77 | 48.20 | 10.43 | 40.00 | 2.23 | 36.77 | -1.00 | 0 | 0.013 | 0.013 |
| | | 11 | 38 | 0.0129 | 0.54 | 4.70 | 4.16 | 0.64 | 0.10 | 0.64 | 0.10 | 1.32 | 0.78 | 35.80 | 42.91 | 7.11 | 39.60 | 3.80 | 70.88 | 35.08 | N/A | | |
| | | 15 | 147 | 0.0302 | 0.18 | N/A | | 0.59 | 0.41 | 0.44 | 0.26 | 0.50 | 0.32 | 11.88 | 35.52 | 23.64 | 30.20 | 18.32 | 29.31 | 17.43 | N/A | | |
| | | 18 | 224 | 0.0380 | 0.32 | 4.16 | 3.84 | 0.51 | 0.19 | 0.34 | 0.02 | 0.72 | 0.40 | 21.00 | 29.22 | 8.22 | 20.40 | -0.60 | 38.60 | 17.60 | 0 | 0.02022 | 0.02022 |
| | | 21 | 241 | 0.0292 | 0.31 | 4.43 | 4.12 | 0.42 | 0.11 | 0.40 | 0.08 | 0.25 | -0.06 | 19.88 | 26.55 | 6.66 | 26.15 | 6.26 | 16.70 | -3.18 | 0.0067 | 0.0313 | 0.0246 |
| | adjuvant | 6 | 115 | 0.0119 | 0.23 | 4.55 | 4.32 | 0.26 | 0.03 | 0.28 | 0.05 | N/A | | 15.40 | 13.80 | -1.60 | 21.80 | 6.40 | N/A | | N/A | | |
| | | 8 | 85 | 0.0136 | 0.18 | 4.96 | 4.78 | 0.60 | 0.42 | 0.45 | 0.27 | | | 13.40 | 30.40 | 17.00 | 26.80 | 13.40 | | | | | |
| | | 10 | 183 | 0.0070 | 0.04 | 4.58 | 4.54 | 0.33 | 0.29 | 0.36 | 0.32 | | | 3.40 | 15.00 | 11.60 | 23.41 | 20.01 | | | | | |
| | | 13 | 130 | 0.0209 | 0.48 | 5.14 | 4.66 | 0.91 | 0.43 | 0.98 | 0.50 | | | 31.75 | 49.00 | 17.25 | 55.56 | 23.81 | | | | | |
| | | 17 | 71 | 0.0166 | 0.36 | 4.55 | 4.19 | 0.51 | 0.16 | 0.27 | -0.09 | | | 19.64 | 30.73 | 11.09 | 17.17 | -2.48 | | | | | |
| | | 22 | 108 | 0.0186 | 0.36 | 0.36 | 4.25 | 3.89 | 0.22 | 0.39 | 0.02 | | | 18.96 | 32.34 | 13.38 | 23.82 | 4.86 | | | | | |
| | n | | 12 | 12 | 12 | 11 | 11 | 12 | 12 | 12 | 12 | 6 | 6 | 12 | 12 | 12 | 12 | 12 | 6 | 6 | 4 | 4 | 4 |
| | Mean | | 132 | 0.021 | 0.35 | 4.34 | 4.36 | 0.85 | 0.23 | 0.48 | 0.14 | 0.78 | 0.36 | 21.45 | 32.36 | 10.91 | 28.30 | 6.85 | 40.77 | 14.97 | 0.0017 | 0.0193 | 0.0176 |
| | SD | | 60 | 0.009 | 0.18 | 1.36 | 0.28 | 0.98 | 0.13 | 0.21 | 0.17 | 0.42 | 0.35 | 10.26 | 11.08 | 6.50 | 11.59 | 10.62 | 18.83 | 14.70 | 0.0034 | 0.0088 | 0.0059 |
| | Min | | 38 | 0.007 | 0.04 | 0.36 | 3.84 | 0.26 | 0.03 | 0.27 | -0.09 | 0.25 | -0.06 | 3.40 | 13.80 | -1.60 | 14.67 | -13.83 | 16.70 | -3.18 | 0.0000 | 0.0125 | 0.0125 |
| | Max | | 241 | 0.038 | 0.66 | 5.22 | 4.78 | 3.89 | 0.43 | 0.98 | 0.50 | 1.32 | 0.78 | 37.77 | 49.00 | 23.64 | 55.56 | 23.81 | 70.88 | 35.08 | 0.0067 | 0.0313 | 0.0246 |
| | CV (%) | | 45.44 | 44.63 | 50.80 | 31.35 | 6.46 | 115.07 | 58.38 | 42.39 | 122.19 | 53.73 | 98.24 | 47.86 | 34.24 | 59.58 | 40.94 | 155.1 | 46.18 | 98.26 | 200.00 | 45.55 | 33.34 |
| Total | Mean | | | | 0.37 | 4.60 | 4.40 | | | | | | | 23.85 | | | | | | | 0.0066 | | |
| | SD | | | | 0.18 | 0.96 | 0.33 | | | | | | | 10.80 | | | | | | | 0.01740 | | |
| | Min | | | | 0.04 | 0.36 | 3.67 | | | | | | | 1.80 | | | | | | | 0.0000 | | |
| | Max | | | | 0.88 | 5.28 | 4.93 | | | | | | | 48.70 | | | | | | | 0.0526 | | |
| | CV (%) | | | | 48.23 | 20.89 | 7.43 | | | | | | | 45.26 | | | | | | | 264.02 | | |

### Appendix E4

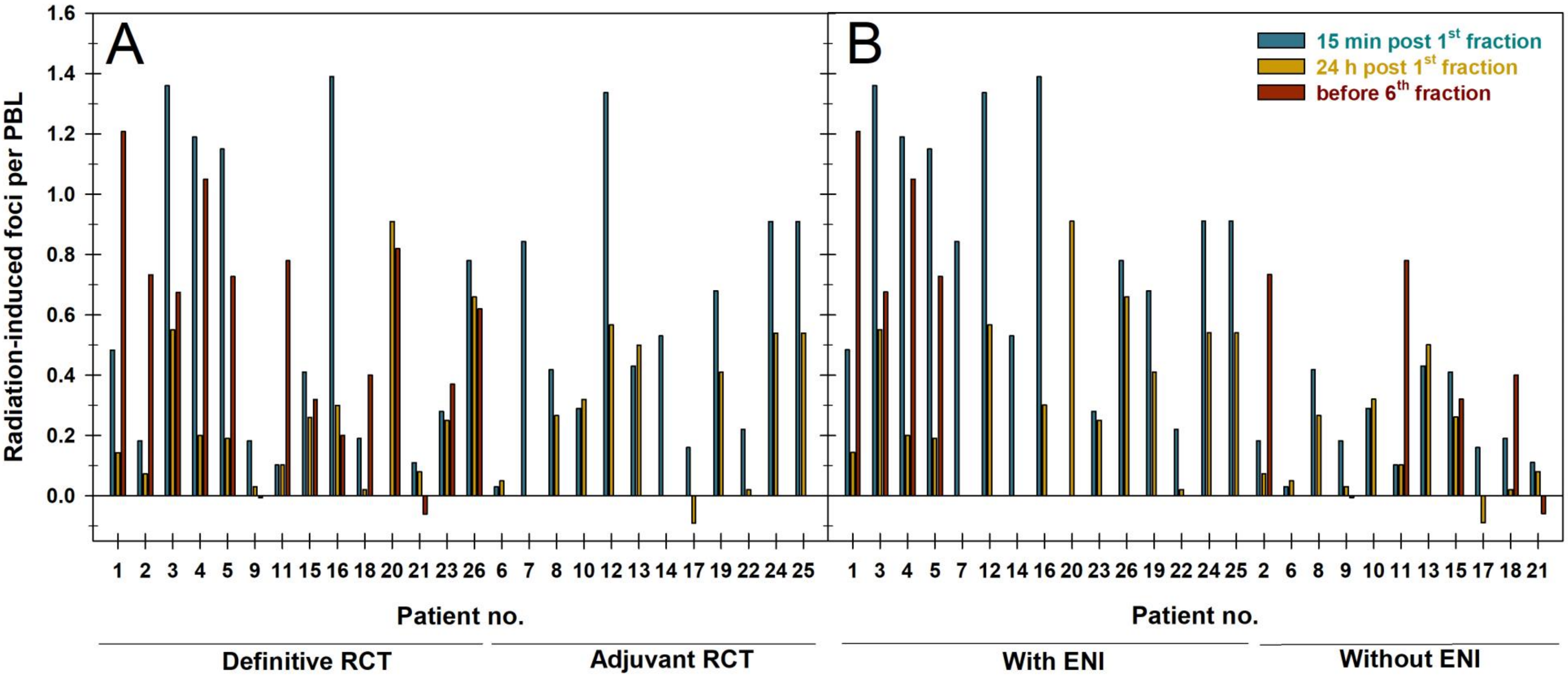


Radiation-induced colocalizing γH2AX/53BP1 foci in peripheral blood lymphocytes (PBLs) post-CRT for all patients stratified according to (A) definitive or adjuvant CRT or (B) elective nodal irradiation status.

### Appendix E5

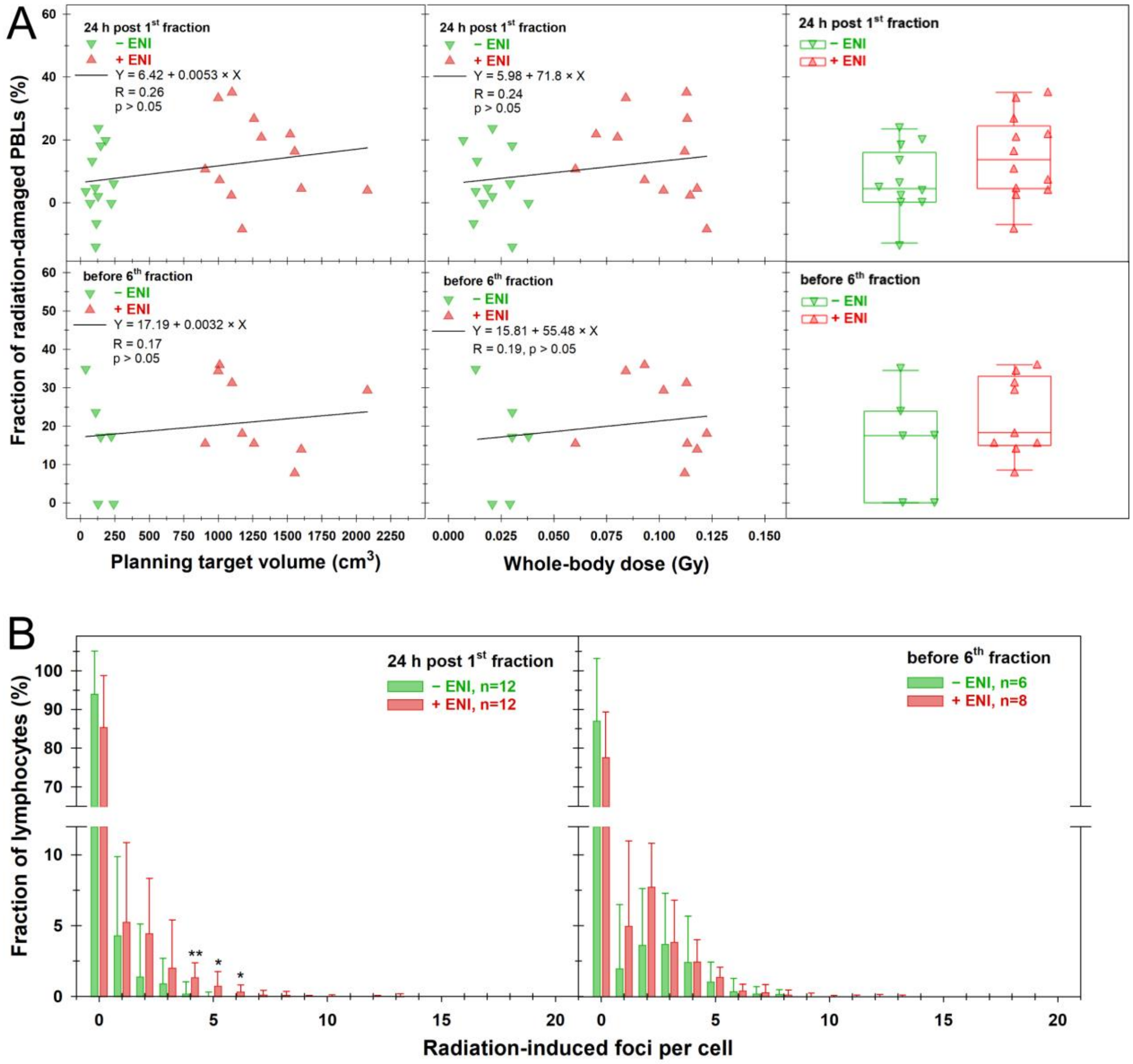


Analysis of the proportion of radiation-damaged peripheral blood lymphocytes (PBLs) and the distribution of biological damage indicators and physical dose distribution in patients. (A) Fraction of PBLs with at least one radiation-induced γH2AX/53BP1 focus after radiotherapy relative to planning target volume (left panel), whole-body dose (middle panel), and stratified by elective nodal irradiation status (± ENI, right panel) 24 h after the first fraction (upper panel) and before the sixth fraction (lower panel). The lines in the left and middle panels represent linear fits to the data, along with the corresponding function, Pearson correlation coefficient (R), and the p-value of the fit. (B) Distribution of radiation-induced γH2AX/53BP1 foci in patients' PBLs 24 h after the first fraction (left panel) and before the sixth fraction (right panel) of radiotherapy ± ENI. Data are presented as the mean and standard deviation. Statistical comparison between radiotherapy ± ENI was conducted using Student's t-test (* $p < 0.05$; ** $p < 0.01$; *** $p < 0.001$).

### Appendix E6

Individual and summarized data for absolute lymphocyte counts per µl collected pre-chemoradiotherapy (CRT) and subsequently every week during CRT and relative lymphocyte counts related to the pre-CRT baseline.

| | | | Absolte lymphocyte counts per µl | | | | | | | | | Relative lymphocyte counts | | | | | | | | |
|---|---|---|---|---|---|---|---|---|---|---|---|---|---|---|---|---|---|---|---|---|
| | | | Weeks after start of CRT | | | | | | | | | Weeks after start of CRT | | | | | | | | |
| Setting | | Pat. no. | 0 | 1 | 2 | 3 | 4 | 5 | 6 | 7 | 8 | 0 | 1 | 2 | 3 | 4 | 5 | 6 | 7 | 8 |
| + ENI | definitive | 1 | 1760 | 1280 | 1210 | 510 | 390 | 370 | N/A | N/A | | 1.000 | 0.727 | 0.688 | 0.290 | 0.222 | 0.210 | | | |
| | | 3 | 1000 | 575 | 260 | 520 | 420 | 180 | 40 | N/A | | 1.000 | 0.575 | 0.260 | 0.520 | 0.420 | 0.180 | 0.040 | N/A | |
| | | 4 | 2270 | 600 | N/A | 600 | 500 | 100 | 200 | 600 | | 1.000 | 0.264 | N/A | 0.264 | 0.220 | 0.044 | 0.088 | 0.264 | |
| | | 5 | 1500 | 1100 | N/A | 500 | 300 | 200 | 200 | 200 | | 1.000 | 0.733 | N/A | 0.333 | 0.200 | 0.133 | 0.133 | 0.133 | |
| | | 16 | 1400 | 600 | 600 | 500 | 300 | 300 | 300 | 300 | | 1.000 | 0.429 | 0.429 | 0.357 | 0.214 | 0.214 | 0.214 | 0.214 | |
| | | 20 | 1300 | 900 | 300 | 700 | N/A | 700 | 600 | 700 | | 1.000 | 0.692 | 0.231 | 0.538 | N/A | 0.538 | 0.462 | 0.538 | |
| | | 23 | 1600 | 500 | 500 | 400 | 400 | 400 | N/A | N/A | | 1.000 | 0.313 | 0.313 | 0.250 | 0.250 | 0.250 | N/A | N/A | |
| | | 26 | 2100 | 2200 | 1100 | 900 | 900 | 600 | 700 | N/A | | 1.000 | 1.048 | 0.524 | 0.429 | 0.429 | 0.286 | 0.333 | N/A | |
| | | 27 | 1600 | N/A | N/A | 300 | 200 | 0 | 400 | 100 | 100 | 1.000 | N/A | N/A | 0.188 | 0.125 | 0.000 | 0.250 | 0.063 | 0.063 |
| | adjuvant | 7 | 1800 | 1400 | 800 | N/A | 600 | 700 | 400 | 500 | | 1.000 | 0.778 | 0.444 | N/A | 0.333 | 0.389 | 0.222 | 0.278 | |
| | | 12 | 2200 | N/A | 1100 | 800 | 1000 | N/A | 300 | N/A | | 1.000 | N/A | 0.500 | 0.364 | 0.455 | N/A | 0.136 | N/A | |
| | | 14 | 1900 | N/A | 2300 | 1600 | N/A | N/A | N/A | N/A | | 1.000 | N/A | 1.211 | 0.842 | N/A | N/A | N/A | N/A | |
| | | 19 | N/A | 1100 | 700 | 1000 | 700 | 500 | 600 | 400 | 500 | N/A | N/A | N/A | N/A | N/A | N/A | N/A | N/A | N/A |
| | | 24 | 1400 | 700 | 400 | 1200 | 1200 | 900 | 1200 | N/A | | 1.000 | 0.500 | 0.286 | 0.857 | 0.857 | 0.643 | 0.857 | N/A | |
| | | 25 | 2600 | 2200 | 1800 | 1200 | 900 | 800 | 800 | N/A | | 1.000 | 0.846 | 0.692 | 0.462 | 0.346 | 0.308 | 0.308 | N/A | |
| | n | | 14 | 12 | 12 | 14 | 13 | 13 | 12 | 7 | 2 | 14 | 11 | 11 | 13 | 12 | 12 | 11 | 6 | 1 |
| | Mean | | 1745 | 1096 | 923 | 766 | 601 | 442 | 478 | 400 | 300 | 1.000 | 0.628 | 0.507 | 0.438 | 0.339 | 0.266 | 0.277 | 0.248 | 0.063 |
| | SD | | 435 | 594 | 622 | 372 | 313 | 284 | 320 | 216 | 283 | 0.000 | 0.237 | 0.282 | 0.210 | 0.194 | 0.187 | 0.227 | 0.164 | |
| | Min | | 1000 | 500 | 260 | 300 | 200 | 0 | 40 | 100 | 100 | 1.000 | 0.264 | 0.231 | 0.188 | 0.125 | 0.000 | 0.040 | 0.063 | 0.063 |
| | Max | | 2600 | 2200 | 2300 | 1600 | 1200 | 900 | 1200 | 700 | 500 | 1.000 | 1.048 | 1.211 | 0.857 | 0.857 | 0.643 | 0.857 | 0.538 | 0.063 |
| | CV (%) | | 24.9 | 54.2 | 67.5 | 48.5 | 52.1 | 64.1 | 66.9 | 54.0 | 94.3 | 0.0 | 37.8 | 55.7 | 48.0 | 57.1 | 70.2 | 82.0 | 65.9 | |
| - ENI | definitive | 2 | 1593 | 1712 | 1120 | 920 | 570 | 440 | 370 | | | 1.000 | 1.075 | 0.703 | 0.578 | 0.358 | 0.276 | 0.232 | | |
| | | 9 | N/A | 1400 | 600 | 500 | 300 | 300 | N/A | | | N/A | N/A | N/A | N/A | N/A | N/A | N/A | | |
| | | 11 | 1700 | 2000 | 600 | 1000 | 600 | 400 | 500 | | | 1.000 | 1.176 | 0.353 | 0.588 | 0.353 | 0.235 | 0.294 | | |
| | | 15 | 1600 | 1700 | 1000 | 500 | 300 | 400 | 200 | 200 | | 1.000 | 1.063 | 0.625 | 0.313 | 0.188 | 0.250 | 0.125 | 0.125 | |
| | | 18 | 1700 | 1300 | 900 | 500 | 400 | 400 | 300 | | | 1.000 | 0.765 | 0.529 | 0.294 | 0.235 | 0.235 | 0.176 | | |
| | | 21 | 1700 | 1200 | 1200 | 700 | 700 | N/A | 300 | 300 | | 1.000 | 0.706 | 0.706 | 0.412 | 0.412 | N/A | 0.176 | 0.176 | |
| | adjuvant | 6 | 2300 | 2100 | 1900 | 1500 | 1100 | 1100 | 700 | 600 | | 1.000 | 0.913 | 0.826 | 0.652 | 0.478 | 0.478 | 0.304 | 0.261 | |
| | | 8 | 1100 | 1000 | 800 | 700 | 300 | 100 | 200 | | | 1.000 | 0.909 | 0.727 | 0.636 | 0.273 | 0.091 | 0.182 | | |
| | | 10 | 1600 | 1800 | 1000 | 700 | 700 | 600 | 400 | | | 1.000 | 1.125 | 0.625 | 0.438 | 0.438 | 0.375 | 0.250 | | |
| | | 13 | 1800 | N/A | 500 | 500 | 300 | N/A | 200 | | | 1.000 | N/A | 0.278 | 0.278 | 0.167 | N/A | 0.111 | | |
| | | 17 | 1000 | 1200 | 600 | 600 | 600 | 500 | N/A | | | 1.000 | 1.200 | 0.600 | 0.600 | 0.600 | 0.500 | N/A | | |
| | | 22 | 1500 | 1000 | 700 | 600 | 500 | 400 | N/A | | | 1.000 | 0.667 | 0.467 | 0.400 | 0.333 | 0.267 | N/A | | |
| | | 28 | 1600 | N/A | N/A | 700 | 400 | N/A | N/A | | | 1.000 | N/A | N/A | 0.438 | 0.250 | N/A | N/A | | |
| | n | | 12 | 11 | 12 | 13 | 13 | 10 | 9 | 3 | | 12 | 10 | 11 | 12 | 12 | 9 | 9 | 3 | |
| | Mean | | 1599 | 1492 | 910 | 725 | 521 | 464 | 352 | 367 | | 1.000 | 0.960 | 0.585 | 0.469 | 0.340 | 0.301 | 0.206 | 0.187 | |
| | SD | | 328 | 389 | 386 | 281 | 231 | 258 | 166 | 208 | | 0.000 | 0.197 | 0.166 | 0.137 | 0.127 | 0.129 | 0.069 | 0.069 | |
| | Min | | 1000 | 1000 | 500 | 500 | 300 | 100 | 200 | 200 | | 1.000 | 0.667 | 0.278 | 0.278 | 0.167 | 0.091 | 0.111 | 0.125 | |
| | Max | | 2300 | 2100 | 1900 | 1500 | 1100 | 1100 | 700 | 600 | | 1.000 | 1.200 | 0.826 | 0.652 | 0.600 | 0.500 | 0.304 | 0.261 | |
| | CV (%) | | 20.5 | 26.1 | 42.4 | 38.8 | 44.3 | 55.7 | 47.1 | 56.8 | | 0.0 | 20.5 | 28.4 | 29.2 | 37.5 | 43.0 | 33.5 | 36.6 | |
